# Highland of COVID-19 outside Hubei: epidemic characteristics, control and projections of Wenzhou, China

**DOI:** 10.1101/2020.02.25.20024398

**Authors:** Liangde Xu, Jian Yuan, Yaru Zhang, Guosi Zhang, Fan Lu, Jianzhong Su, Jia Qu

**Affiliations:** Institute of Biomedical Big Data, Wenzhou Medical University, Wenzhou, 325027, China; School of Ophthalmology & Optometry and Eye Hospital, Wenzhou Medical University, Wenzhou, 325027, China; State Key Laboratory of Ophthalmology, Optometry and Visual Science, Wenzhou, 325027, China

**Author notes:** These authors contributed equally to the paper as first authors (Liangde Xu, Jian Yuan). To whom correspondence should be addressed. Corresponding Authors: Jia Qu, M.D.; Jianzhong Su Ph.D., Institute of Biomedical Big Data, School of Ophthalmology & Optometry and Eye Hospital, State Key Laboratory of Ophthalmology, Optometry and Visual Science, Wenzhou Medical University, Xueyuan road #270, Wenzhou 325027, China.

## Abstract

In late December 2019, Chinese authorities reported a cluster of pneumonia cases of unknown aetiology in Wuhan, China^1^. A novel strain of coronavirus named Severe acute respiratory syndrome coronavirus 2 (SARS-CoV-2) was isolated and identified on 2 January 2020 ^2^. Human-to-human transmission have been confirmed by a study of a family cluster and have occurred in health-care workers ^3,4^. Until 10 February 2020, 42638 cases of 2019 novel coronavirus disease (COVID-19) have been confirmed in China, of which 31728 came from Hubei Province (Figure). Wenzhou, as a southeast coastal city with the most cases outside Hubei Province, its policy control and epidemic projections have certain references for national and worldwide epidemic prevention and control. We described the epidemiologic characteristics of COVID-19 in Wenzhou and made a transmission model to predict the expected number of cases in the coming days.

## Methods

In this epidemiologic and modelling study, we first gathered transmission data for 434 confirmed COVID-19 patients in the ongoing Wenzhou outbreak from the the National Health Commission of the People’s Republic of China and the Health Commission of Wenzhou by February 10, 2020. We then investigate a deterministic (susceptible-exposed-infectious-recovered) SEIR compartmental model based on the clinical progression of the disease, epidemiological status of the individuals, and the intervention measures to inferred the basic reproductive number of SARS-CoV-2 and the transmission among Wenzhou and three prefecture-level cities. Finally, we forecasted and compared the spread of SARS-CoV-2 across Wenzhou (Southeastern), Shenzhen (Southern), Zhengzhou (Central) and Harbin (Northern), accounting for the effect of the Wenzhou quarantine implemented since Jan 27, 2020 and other interventions. See https://github.com/ZhangBuDiu/WZ_COVID-19 for R code, case data, and prepared datafiles.

## Results

A total of 434 incidence, 31 severe case, 99 hospital discharges and 5304 probable COVID-19 cases were reported before February 10, 2020, and 168 patients who visited Wuhan after the onset of the SARS-CoV-2 epidemic. The median age of persons with COVID-19 was 47 years; 48 cases (11.1%) occurred in persons 65 years of age or older, and 5 (1.2%) were in children younger than 15 years of age (Table). The maximum number of illness onset in Wenzhou appeared on January 26 (38 cases). Since January 27, the incidence per day in Wenzhou has continued to decline is likely to be due to isolation of people return to Wenzhou, implementation of region quarantine and suspension of public transportation system. Under the current control efforts, we estimate that the daily incidence by March 3-9, 2020 will drop to 0 in Wenzhou using SEIR model. The total number of affected people is 538 (95% CI: 534-542). Estimated of the basic reproduction numbers, R0, were 2.91 (95% CI: 2.35-3.57) for Wenzhou, 2.53 (95% CI, 1.86 to 3.34) for Shenzhen, 5.95 (95% CI, 5.36 to 6.67) for Zhengzhou, and 3.35 (95% CI, 3.07 to 3.68) for Harbin. Figure shows the trajectories of the epidemic for Wenzhou, Shenzhen, Zhengzhou and Harbin. We classified these cities to Input-Burst, Input-Control, Control-Relaxation and Control-Missing based on epidemic dynamics and control measures. We found that the epidemic in Wenzhou was composed of outsiders (people with a visit history to Hubei) and local people, while the epidemic in Shenzhen was mainly composed of outsiders. Under strict implemented control measures, epidemic gradually subsided in both Wenzhou and Shenzhen.

**Figure.**
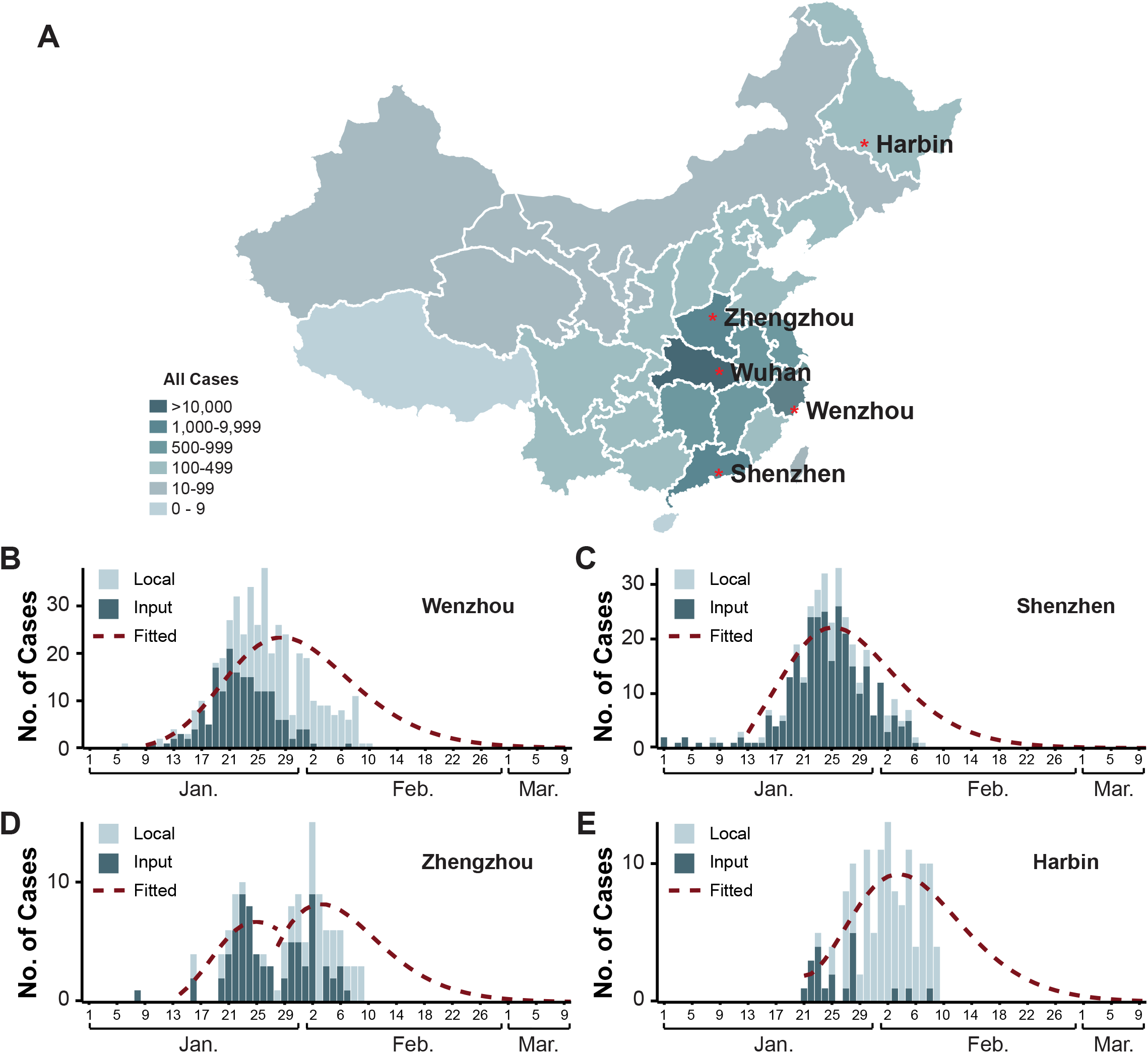
Comparison of observed cases and predicted detected cases for Wenzhou and three Chinese cities. Panel A. Geographic Distribution of SARS-CoV-2 Infection in China, as of February 10, 2020. Panel B through E show the observed (histograms) and projected (dashed line) daily case incidence for four cities, Wenzhou (B), Shenzhen (C), Zhengzhou (D) and Harbin (E), respectively.

**Table.**
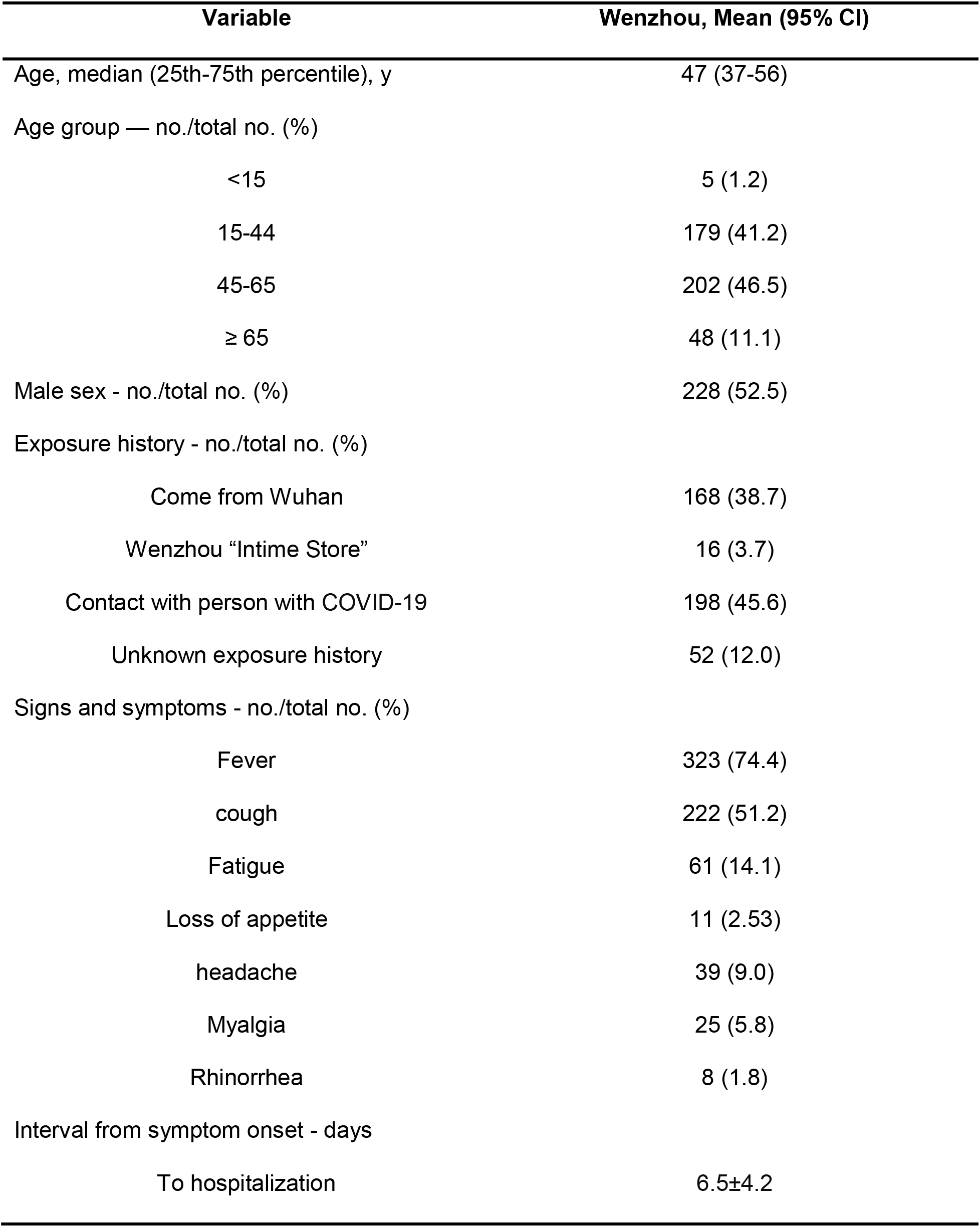
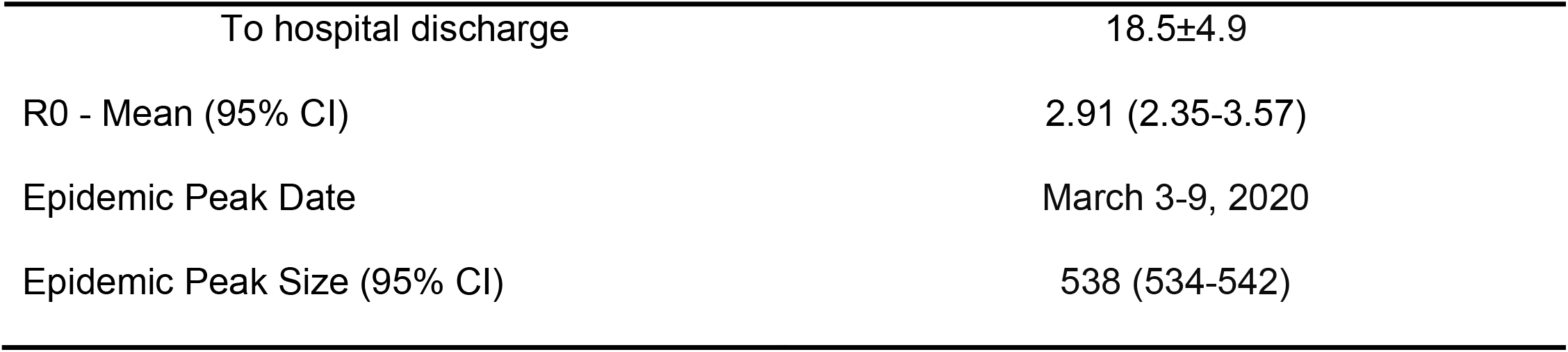
Epidemiologic Characteristics and Variables of Patients with COVID-19 in Wenzhou as of February 10, 2019.

| Variable | Wenzhou, Mean (95% CI) |
| --- | --- |
| Age, median (25th-75th percentile), y | 47 (37-56) |
| Age group — no./total no. (%) |  |
| <15 | 5 (1.2) |
| 15-44 | 179 (41.2) |
| 45-65 | 202 (46.5) |
| ≥ 65 | 48 (11.1) |
| Male sex - no./total no. (%) | 228 (52.5) |
| Exposure history - no./total no. (%) |  |
| Come from Wuhan | 168 (38.7) |
| Wenzhou “Intime Store” | 16 (3.7) |
| Contact with person with COVID-19 | 198 (45.6) |
| Unknown exposure history | 52 (12.0) |
| Signs and symptoms - no./total no. (%) |  |
| Fever | 323 (74.4) |
| cough | 222 (51.2) |
| Fatigue | 61 (14.1) |
| Loss of appetite | 11 (2.53) |
| headache | 39 (9.0) |
| Myalgia | 25 (5.8) |
| Rhinorrhea | 8 (1.8) |
| Interval from symptom onset - days |  |
| To hospitalization | 6.5±4.2 |
| To hospital discharge | 18.5±4.9 |
| R0 - Mean (95% CI) | 2.91 (2.35-3.57) |
| Epidemic Peak Date | March 3-9, 2020 |
| Epidemic Peak Size (95% CI) | 538 (534-542) |

## Discussion

Although the current epidemic of SARS-CoV-2 in China is unprecedented in scale, clinical presentations greatly resemble SARS-CoV. The higher number of infections may be due to the late identification of pathogens, the host’s ability to hide infection symptoms and high transport capacity of China. As we known, estimate of epidemic dynamics and predictions are of crucial importance for public health planning and control measurement ^5,6^. In this study, we provide an assessment of the transmission dynamics and predictions of the future spread of COVID-19 among four cities. Forward projections suggest that the Wenzhou and Shenzhen epidemic will end before Zhengzhou and Harbin, which result from substantial and draconian measures that limit population movements and drastically reduce within-population contact rates, such as greater community engagement, cancellation group gatherings, school closures, and work-from-home arrangements.

## Data Availability

https://github.com/ZhangBuDiu/WZ_COVID-19 for R code, case data, and prepared datafiles

## Author Contributors

J. Qu, J. Su and L. Xu conceived and designed the study; J. Yuan, L. Xu G. Zhang, R. Zhang and Y. Huang collected and analysed data; J. Yuan, J. Su, L. Xu and J Qu drafted the manuscript.

## References

1. Tan W, Zhao X, Ma X, Wang W, Niu P, Xu W. A novel coronavirus genome identified in a cluster of pneumonia cases—Wuhan, China 2019− 2020. China CDC Weekly. 2020;2(4):61–62.

2. Zhu N, Zhang D, Wang W, et al. A novel coronavirus from patients with pneumonia in China, 2019. New England Journal of Medicine. 2020.

3. Wang D, Hu B, Hu C, et al. Clinical Characteristics of 138 Hospitalized Patients With 2019 Novel Coronavirus–Infected Pneumonia in Wuhan, China. JAMA. 2020.

4. Chan JF-W, Yuan S, Kok K-H, et al. A familial cluster of pneumonia associated with the 2019 novel coronavirus indicating person-to-person transmission: a study of a family cluster. The Lancet. 2020.

5. Wu JT, Leung K, Leung GM. Nowcasting and forecasting the potential domestic and international spread of the 2019-nCoV outbreak originating in Wuhan, China: a modelling study. The Lancet. 2020.

6. Li Q, Guan X, Wu P, et al. Early Transmission Dynamics in Wuhan, China, of Novel Coronavirus–Infected Pneumonia. New England Journal of Medicine. 2020.

